# External control arm analysis: an evaluation of propensity score approaches, G-computation, and doubly debiased machine learning

**DOI:** 10.1101/2022.01.28.22269591

**Authors:** Nicolas Loiseau, Paul Trichelair, Maxime He, Mathieu Andreux, Mikhail Zaslavskiy, Gilles Wainrib, Michael G.B. Blum

**Author notes:** Correspondence Owkin France, Paris, France. Equal contributor (Nicolas Loiseau, Paul Trichelair).

## Abstract

**Background:** An external control arm is a cohort of control patients that are collected from data external to a single-arm trial. To provide an unbiased estimation of efficacy, the clinical profiles of patients from single and external arms should be aligned, typically using propensity score approaches. There are alternative approaches to infer efficacy based on comparisons between outcomes of single-arm patients and machine-learning predictions of control patient outcomes. These methods include G-computation and Doubly Debiased Machine Learning (DDML) and their evaluation for ECA analysis is insufficient.

**Methods:** We consider both numerical simulations and a trial replication procedure to evaluate the different statistical approaches: propensity score matching, Inverse Probability of Treatment Weighting (IPTW), G-computation, and DDML. The replication study relies on five type 2 diabetes randomized clinical trials granted by the Yale University Open Data Access (YODA) project. From the pool of five trials, observational experiments are artificially built by replacing a control arm from one trial by an arm originating from another trial and containing similarly-treated patients.

**Results:** Among the different statistical approaches, numerical simulations show that DDML has the smallest bias followed by G-computation. Ranking based on mean square error is different with G-computation always being among the lowest-error methods while DDML relative performance improves with increasing sample sizes. For hypothesis testing, DDML controls type-1 error and is conservative whereas G-computation and propensity score approaches can be liberal with type I errors ranging between 5% and 10% in some settings. G-computation is the best method in terms of statistical power, and DDML has comparable power at *n* = 1000 but its power is inferior to propensity score approaches at *n* = 250. The replication procedure also indicates that G-computation minimizes mean squared error while DDML has intermediate performances compared to G-computation and propensity score approaches. The confidence intervals of G-computation are the narrowest in lines with its liberal type I error whereas confidence intervals of DDML are the widest that confirms its conservative nature.

**Conclusions:** For external control arm analyses, methods based on outcome prediction models can reduce estimation error and increase statistical power compared to propensity score approaches.

## Background

There is an increasing interest in using external control arms (ECA) as a source of evidence to assess treatment efficacy. An ECA consists of a cohort of patients that serve as controls to an intervention arm from a clinical trial, and these control patients are collected from data sources external to the single-arm trial [1, 2]. After running a single-arm phase 2 study, usage of ECA is relevant to reduce false positive rates [3]. ECAs are also relevant to supplement randomized trials when randomization is unethical or when it is difficult to recruit patients, typically for rare diseases or in precision oncology where recruitment relies on biomarkers [4].

However, causal inference in non-randomized studies such as ECA is prone to confounding bias [5, 6]. Without randomization, estimation of treatment effect can be biased partly because of differences between the characteristics of patients in the two arms. Methods based on propensity scores are well established to account for confounding factors [7–10]. Propensity scores relies on the exposure model that provides a mapping between patient characteristics and the probability to be in the external arm. As an alternative, there are several methods that require prediction of clinical outcomes based on covariates and on treatment [11, 12]. In epidemiology, G-computation is such an alternative, and it is based on the *counterfactual framework* in which we posit that we can predict a patient outcome if the patient would have been enrolled in the control arm instead of the experimental one or vice-versa, making the inference of a causal effect theoretically possible [11]. With the advent of causal inference in machine learning, the counterfactual framework has been reinvestigated and new methods were proposed including doubly debiased machine learning [12], which addresses bias of machine learning estimators. Here, we consider both synthetic simulations and data of clinical trials to evaluate statistical properties of both propensity score and outcome prediction methods. Evaluated methods seek to estimate the average treatment effect on the treated (ATT), which is defined as the benefit of the investigated treatment when averaged over the characteristics of the individuals originating from the intervention arm of the clinical trial.

The first class of statistical methods relies on propensity scores that are computed after learning an exposure model *e*, which relates individual covariates to the probability to lie in the experimental arm. Exposure model can be estimated using a logistic regression. Treatment effect is then estimated using patients matching and/or weighting, such as the distribution of the propensity scores should be the same in both arms. Rosenbaum an Rubin [13] showed that if positivity and conditional ignorability hold, then conditioning on the propensity score allows to obtain unbiased estimates of average treatment effects [14]. Conditional ignorability means that there are no unmeasured confounders. Mathematically, it states that given a set of covariates *X*, treatment assignment *T* is independent of the potential outcomes (*Y* ^0^, *Y* ^1^) that would be realized when the treatment *T* is equal to 0 (control) and 1 (investigated treatment). The second assumption is positivity and it assumes that 0 < *P* (*T* = 1|*X*) < 1, for all values of *X*, which means that every subject has a nonzero probability to receive the control treatment and the investigated treatment. If the exposure model is misspecified, potentially because parametric assumptions of logistic regression are not valid, then estimators of treatment effect might be biased [15].

The second class of methods, outcome prediction methods, relies on the outcome model *μ*_0_, sometimes named Q-model, which is the conditional expectation of the clinical outcome based on covariates *X* [11]. Because we focus on the estimation of the average treatment effect on the treated (ATT), the nuisance function *μ*_0_ corresponds to the expected outcome for a patient enrolled in the control arm (see Methods). By contrast, estimation of the average treatment effect (ATE) would have required outcome prediction as function of both the treatment and the covariates, which is the standard definition of the Q model [11]. Fitting the Q model can be done with flexible machine learning models such as boosted trees or neural networks [16, 17]. Machine learning models can be trained using regularization to limit overfitting. However, while reducing variance of estimators, regularization can bias estimation of outcome model that can in turn bias estimation of treatment effect [12]. Doubly debiased machine learning (DDML) is related to G-computation but it further accounts for the possible bias of machine learning outcome models [12]. DDML requires to estimate both the exposure model *e* and the outcome model *μ*_0_, and flexible models can be fitted to infer both *e* and *μ*_0_, which are considered as nuisance parameters [18]. To provide unbiased estimation of treatment effect, DDML relies on Neyman orthogonal scores and on cross fitting, which is a sample splitting approach [12].

There is a lack of studies based on clinical trial data that compares propensity score approaches and methods based on outcome modelling. Numerical simulations suggest that G-computation reduces bias and variance of causal inference estimate compared to propensity-score approaches [19, 20]. Another simulation study finds that DDML was among the top performers methods to estimate average treatment effect [21]. However, comparisons based on actual trial data are insufficient. Here we consider an internal replication framework for evaluation of causal inference methods [22]. It is based on comparisons between randomized studies that provide ground truths for treatment effect and artificial non-randomized studies consisting of the grouping of the experimental arm and of the standard-of-care arm, which are derived from two different clinical trials [23]. An internal replication framework was used for instance to demonstrate that propensity score matching is highly sensitive to baseline covariates included in the exposure model [24]. Internal replication framework are not the only setting to compare results from RCT and from observational data. Several studies compared results obtained from observational data to the conclusions obtained from randomized experiments, which are considered as ground truth [25–28]. However, heterogeneity of treatment effect can explain the difference of efficacy measured in a RCT and observational setting [29, 30]. By contrast, there is no expected difference of treatment effect (ATT) in internal replication studies when comparing efficacy obtained from randomized and non-randomized experiment [22]. Our internal replication study is based on data from the YODA project, which includes a pool of type-2 diabetes randomized clinical trials sharing arms with the same treatment delivered to patients (Canagliflozin) [31, 32].

## Methods

### Average Treatment effect on the Treated (ATT)

Generally, the primary quantity of interest in interventional clinical trials is the efficacy of an investigated treatment compared to another standard of care or placebo treatment. Formally, from the study cohort comprising of two groups, each exposed to a different treatment T (0 for control, 1 for experimental treatment), the target is to infer the average treatment effect on the treated (ATT). The ATT corresponds to the difference between the outcome of a patient treated with the experimental drug and a control patient when averaging over baseline clinical attributes *X* of patients belonging to the experimental treatment arm. Using the formalism of potential outcomes, the ATT is defined as [33]

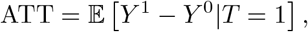

where *Y* ^0^ (respectively *Y* ^1^) is the potential outcome for a unit that undergoes treatment 0 (respectively 1). The observed outcome *Y* can be expressed as

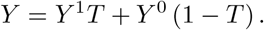

For a given patient, only one of the two potential outcomes is realized and observed, the other is named a counterfactual outcome. The ATT estimand is different from the average treatment effect that is obtained as

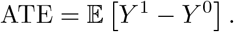

If either the propensity score *e*(*X*) = 𝔼 [*T*|*X*] is constant (randomization) or the Conditional Average Treatment Effect CATE = 𝔼[*Y*^1^ − *Y*^0^|*X*] is constant (no heterogeneity), then ATT and ATE are equal. In the following, we will also denote by *μ*_0_ = 𝔼 [*Y*^0^|*X*], the conditional expectation of the outcome for patients in the control arm.

### Estimators of the average treatment effect of the treated

The problem of causal inference for external control arm analysis revolves around the two populations’ prognosis characteristics not being of equal distribution in the two arms. A solution to balance populations’ characteristics is to reweight or choose units such that the two resulting virtual populations match as closely as possible. To balance populations, the exposure model *e*(·) should be estimated when considering propensity score matching (PSM) and Inverse Probability of Treatment Weighting (IPTW). The PSM estimator selects matched units in each group whereas IPTW re-weights units based on functions of the propensity score, which leads to the following estimator [34, 35]

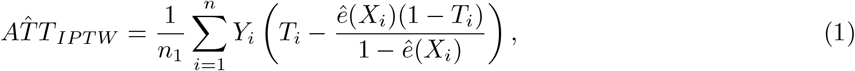

where *X*_*i*_, *Y*_*i*_, *T*_*i*_ are the covariates, outcome, and treatment for the *i*^*th*^ individual, 1, …, *n*_1_ are the indices of the individuals in the experimental arm, *n*_1_+ 1,…, *n* are the indices of the individuals in the external arm, *n, n*_1_ are the sample sizes for the whole sample and the experimental arm only, and *ê* is an estimator of the exposure model.

The first estimator based on outcome prediction we consider is the G-computation estimator [19, 36]. G-computation does not rely on estimation of the propensity score but on the the conditional expectation of the outcome *μ*_0_. For each treated patient defined by his covariates *X* and outcome *Y*, we can predict a control counter-factual outcome 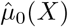, and the *G*-computation estimator is defined as the average over the experimental arm of the difference between the measured and counterfactual outcome

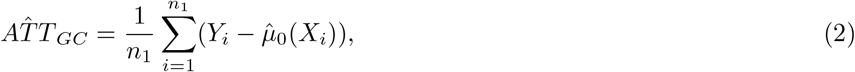

where 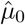 is an estimator of the nuisance function.

Machine learning estimators can be biased in order to avoid overfitting and this is especially true when the dimension of the covariates *X* is large [37]. Doubly debiased machine learning (DDML) accounts for the bias of the G-computation estimator, which can result from the bias of a machine learning estimator, 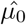, for *μ*_0_ [12]. A core principle of DDML is to consider a sample splitting approach to estimate and account for the bias of the machine learning estimator of the outcome model. The dataset is split into a training set and an auxiliary set. The training set is used to fit two machine learning models to learn the outcome and exposure models *μ*_0_ and *e*. The ATT estimator is obtained by subtracting to the G-computation estimator evaluated on the auxiliary dataset an estimate of its bias

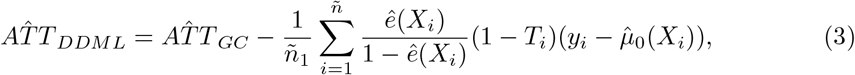

where *ñ, ñ*_1_ are the sample sizes for the whole auxiliary dataset and the control arm part of this dataset. Because the estimator depends on actual splitting, we consider an averaging procedure over multiple splits [12]. In the Appendix, we describe the averaging procedure and the estimation procedure for the variance.

Finally, we compute an unadjusted estimator that consists of the difference between the mean of the clinical outcomes *Y* in each arm. This estimator measures the level of bias that is expected when not accounting for confounding factors. If the data include confounding that may impact causal inference, the unadjusted estimator should be biased.

### Variance, confidence intervals, and regularisation

To estimate variance and confidence intervals we consider non-parametric bootstrap with 100 replicates for both the propensity score approach and G-computation. For G-computation, the bootstrap procedure was applied before fitting the *μ*_0_ function. For DDML, we consider a sample-splitting approach [12]; estimation of the variance is detailed in the Appendix.

For all methods, we consider linear regression and logistic regression with all covariates to fit *μ*_0_ and *e*. To train the propensity score model *e*, we consider ridge regression, and to train the outcome model *μ*_0_, we consider lasso regression. For G-computation, regularisation parameters were learned using cross-validation. For DDML, regularisation parameters were learned using nested cross-validation because of the internal cross-validation procedure described in the Appendix. Machine learning operations were performed using the *Scikit-learn* Python library [38].

### Synthetic Simulations

We consider two scenarios of simulations to benchmark estimators. The first scenario assumes an homogeneous treatment effect and includes confounding factors because both the exposure and the outcome models are linear functions of several of the 20 simulated covariates. The second scenario further assumes an heterogeneous treatment effect by including interaction between treatment and covariates to model outcomes.

Experiments are based on synthetic data with a binary exposure *T* and 20 covariates *X*. The numbers of patients (including patients in both arms) of 250, 500 and 1000, were chosen to be in the same order of magnitude as external control arm analyses. The simulations rely on two scenarios differing by the potential outcomes (*Y* ^0^, *Y* ^1^) generation. For both scenarios, the exposure model is a linear function of 5 of the 20 covariates.

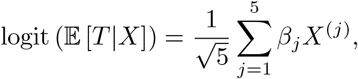

where *β*_*j*_ ∼ 𝒰([−1, 1]), *X* ∼ 𝒩(0, Σ) with Σ a random sparse symmetric definite positive matrix, and where *X*^(*j*)^ is the *j*^*th*^ element of the vector of covariates *X*. In the first scenario, the potential outcomes are sparse linear functions of the covariates and the treatment effect is homogeneous among the patients. To make it sparse, half of the variables are randomly sampled and the corresponding coefficient is set to zero,

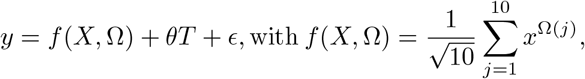

where *ϵ* ∼ *𝒩*(0, 1), Ω is a random permutation of the covariate indices and *θ* ∼ 𝒩(0, 0.4) or *θ* = 0 for the null hypothesis. We chose a variance of 0.4 because we have found in simulations that this value induces a level of confounding that biases the unadjusted estimator, and which can be handled with causal inference approaches.

The second scenario includes a term of interactions to model an heterogeneity of treatment. The outcome is obtained as follows :

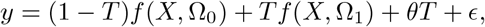

where *ϵ* ∼ 𝒩(0, 1), Ω_1_, Ω_2_ are random permutations of the covariate indices, and *θ* is sampled such that ATT ∼ 𝒩(0, 0.4) or ATT = 0.

To evaluate the estimators, the following metrics were considered : bias, mean absolute error (MAE), mean squared error (MSE), average confidence interval length measured by the variance of a matched Gaussian distribution, type I error and power.

### Internal replication study

The internal replication study is based on data from five randomized clinical trials assessing the efficacy of Canagliflozin in patients with type 2 diabetes [39–43]. Access to the trials, shortly described in Table 1, was granted through the Yale University Open Data Access (YODA) Project [31, 32]. Experiments are restricted to the set of patients that share similar background therapy and inclusion/exclusion criteria in order to make causal inference valid because of the positivity assumption. A set of 40 baseline covariates were selected by a clinician and considered as confounding factors (see Appendix). The primary endpoint is change in HbA1c (glycated hemoglobin) between baseline and 12 weeks, which is available in all trials. Patients with missing outcome are not considered in the analysis.

**Table 1.**
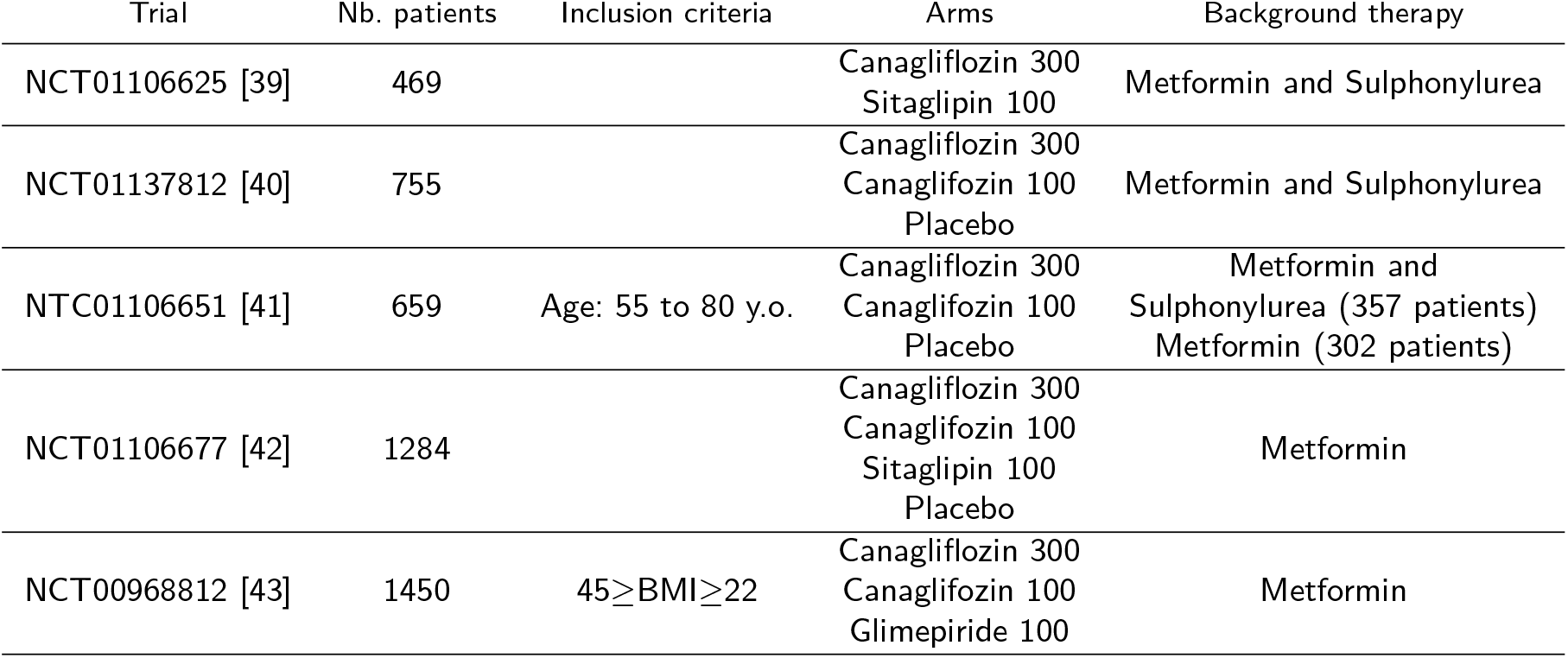
Description of the five type 2 diabetes clinical trials used for the internal replication study. We report only the trial-specific inclusion criteria.

From the pool of five trials, an observational setting is built by replacing a control arm in one trial by another trial arm composed of patients that were given the same treatment. This procedure is replicated by varying the trial of interest. Estimation obtained in the non-randomized setting can be compared to the treatment effect obtained in the well randomized setting.

We conduct two categories of internal replication studies. For each experiment, the experimental arm and the control arm are extracted from different trials. In the first category, the experimental and control treatments are the same. In this negative control setting, the treatment effect on the treated is null regardless of the underlying population [44]. The negative control study is based on 9 non-randomized comparisons. The ground truth of a null effect being known, the comparison between the estimators is performed using the following metrics: the mean absolute error (MAE), the mean squared error (MSE), the width of confidence intervals, and the coverage rates for the 95% confidence intervals.

In the second category of experiments, the experimental and control treatments are different and an RCT estimate is available from one of the five trials listed in Table 1. In this RCT replication setting, a reference treatment effect and confidence intervals are available from the RCT but the true treatment effect is unknown. The RCT replication study is based on 19 non-randomized comparisons. Evaluation relies on previously proposed metrics [45]:

- Pseudo bias is defined as the difference between the randomized treatment effect estimation and the non-randomized estimation;
- Pseudo mean squared error is defined as the squared difference between the randomized effect estimation and the non-randomized estimation, averaged over the different combinations of trials;
- Estimate agreement measures the percentage of time when treatment effect estimated in the non-randomized setting lies within the 95% confidence interval of the randomized trial;
- Regulatory agreement is the percentage of time the cutoff *P* < 0.05 obtained with the non-randomized experiments agrees with the RCT result about *P* < 0.05.

## Results

### Synthetic Simulations

Type I error rates and statistical power are evaluated with simulations using *P* < 0.05 as a decision cutoff. The unadjusted estimator has an inflated type I error ranging from 10 − 20% when *n* = 250 to 30 − 45% when *n* = 1000 showing that simulations include a confounding bias (Figure 1). The statistical power obtained with the unadjusted estimator is of poor relevance because of its inflated type I error. Among the methods that adjust for confounding bias, we find a slight excess of type I error for propensity-score methods and G-computation with type I error values that range between 5% and 10%. The propensity score approaches have type I error rates that range between 0% and 10%. The 10% error rate is reached when *n* = 1, 000 and when simulations include interaction terms between treatment and covariates. The DDML method has the lowest type I error that stays below or at the order of the 5% nominal threshold.

**Figure 1.**
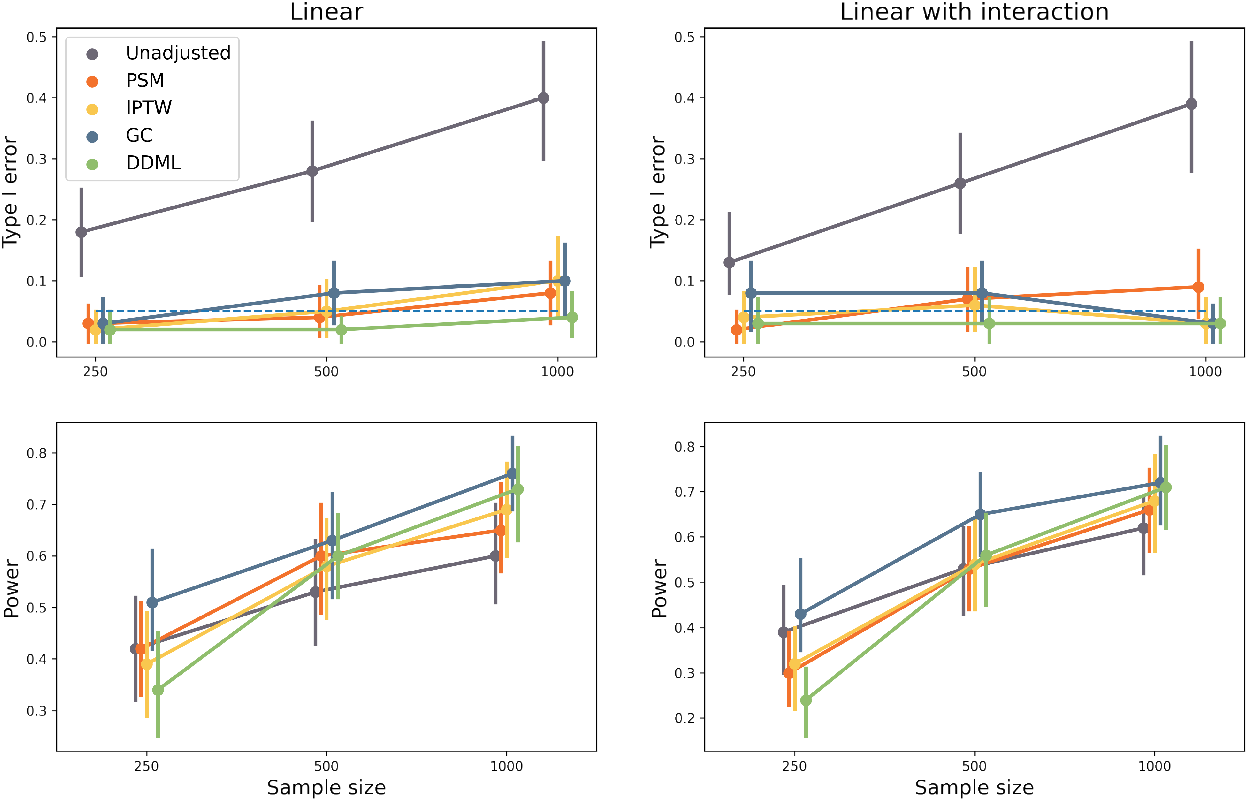
Type I error rate and power evaluated with Monte Carlo simulations of the five estimators included in the simulation study. Each dot corresponds to a simulation study that includes 100 replicates. The horizontal dashed line corresponds to the expected type I error rate of 5%.

When considering G-computation, power is increased by 0 − 10% when compared to propensity-score approaches (Figure 1). By contrast, the power of DDML is not always larger than the ones of propensity-score approaches. The power of DDML is smaller that the ones obtained with propensity score approaches when *n* = 250, of comparable values at *n* = 500, and larger when *n* = 1000. As expected, the power of each method increases with increasing sample size.

Statistical properties of the different estimators are also compared using Mean Absolute Error (MAE) and the Mean Squared Error (MSE). Both errors indicate that there are three groups of estimators as ranked by their performances (Figure 2). The unadjusted difference of means provides the worst estimate of ATT. Then, the second group includes the two propensity score approaches. They have similar performances than methods of the third group based on outcome modelling (G-computation and DDML) for small sample size at *n* = 250. However, their errors decrease more slowly than outcome modelling methods resulting in larger errors especially when *n* = 1000. When comparing outcome modelling methods, G-computation has a lowest error compared to DDML except when *n* = 1, 000 for the Monte Carlo simulations without treatment effect heterogeneity.

**Figure 2.**
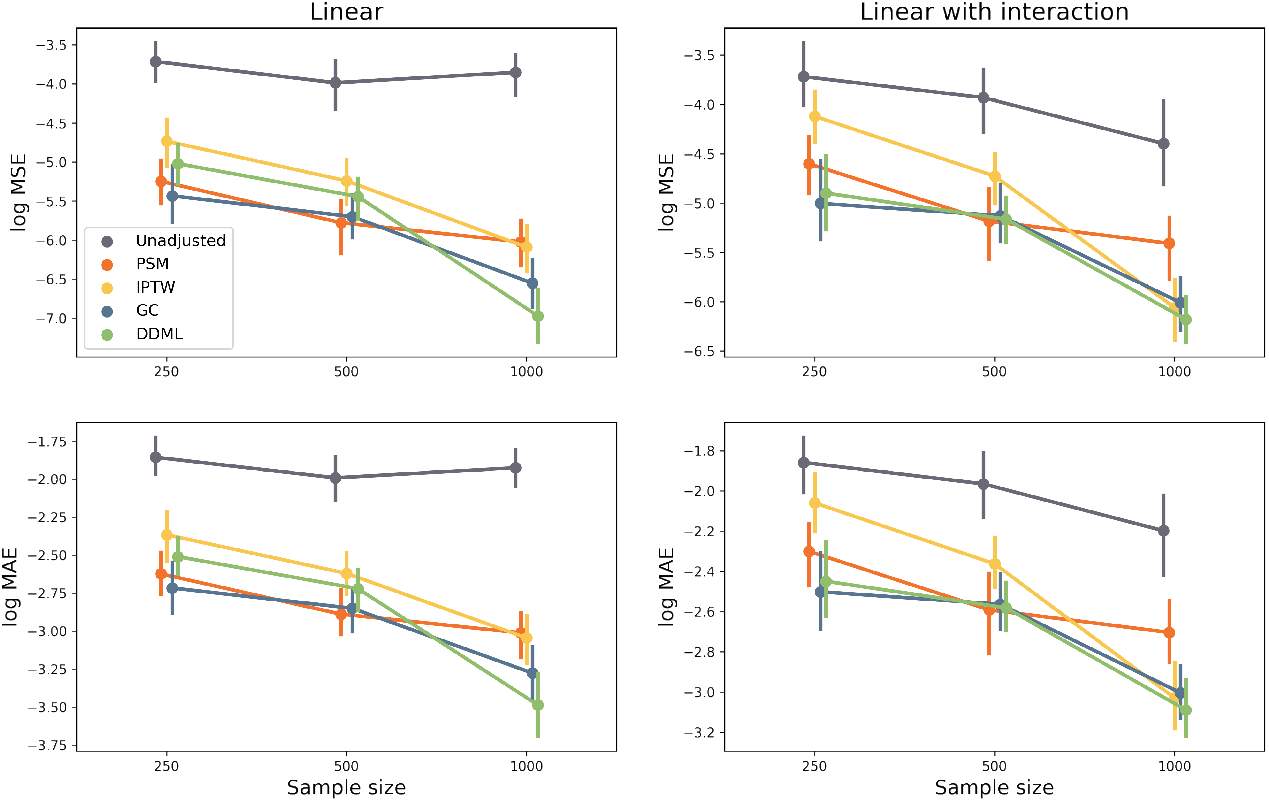
Logarithm of the Mean Absolute Error (MAE) and Mean Squared Error (MSE) of the five estimators included in the simulation study. Each dot corresponds to a simulation study that includes 100 replicates.

To have a finer look at the different properties of ATT estimators, we investigate their bias (Figure 3). As expected by construction of the DDML estimator, its bias is inferior to the bias of G-computation. For this outcome prediction methods, bias decreases a a function of sample size. The bias of propensity score methods was larger than the ones of outcome prediction methods and it does not monotonically decrease with sample size.

**Figure 3.**
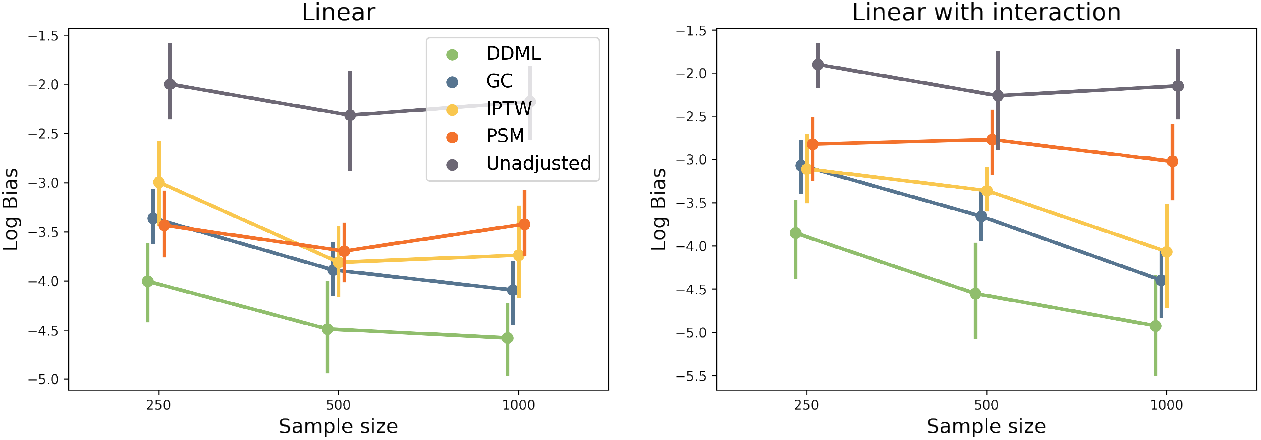
Logarithm of the bias of the five estimators included in the simulation study. Each dot corresponds to a simulation study that includes 100 replicates.

Last, we capture the width of the confidence intervals by computing the variance of a Gaussian distribution which 95% C.I. match the observed 95% C.I. width (Figure 4). G-computation produces the narrowest credibility intervals and as expected their width decreases with increasing sample sizes. The width decrease is more pronounced for DDML. At *n* = 250, DDML produces the widest confidence intervals of all methods whereas for *n* = 1, 000, its C.I. width is inferior or comparable to the ones obtained with propensity score methods.

**Figure 4.**
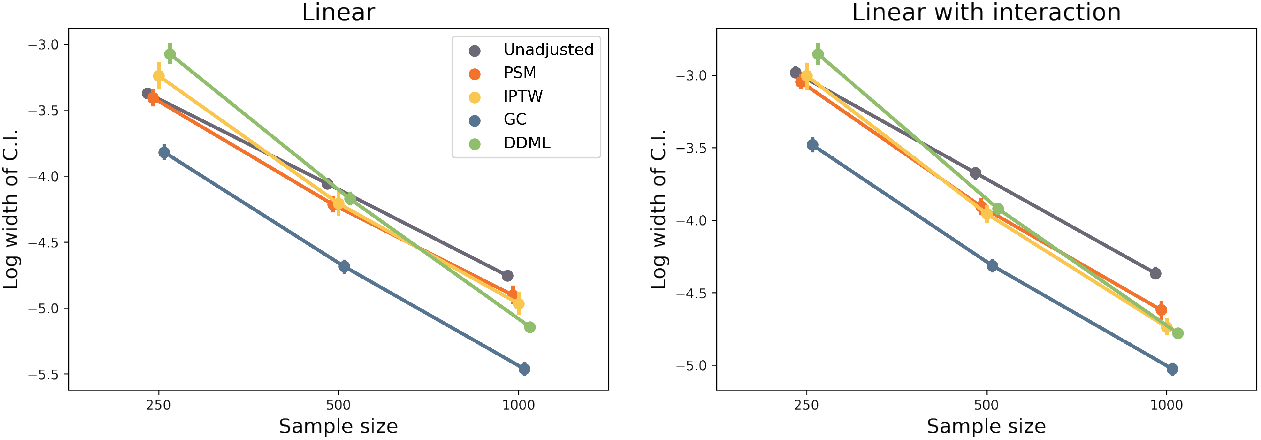
Log width of the 95% Credibility Intervals (C.I) for the different methods. To measure the log width, we compute the logarithm of the variance of a Gaussian distribution which 95% C.I. would match the observed C.I.

### Internal replication study

The internal replication study confirms simulation results. G-computation has the smallest MSE and MAE errors for both null and trial replication (Figure 5, Tables 2 and 3). By contrast the unadjusted approach has the worst performance in terms of MAE and MSE. The two propensity score methods and DDML have intermediate performances (Tables 2 and 3). For null replication, DDML has better performance than IPTW, and PSM has the worst performance (Table 2). For trial replication, DDML has better performance than IPTW, and PSM has the better or worst performance of the three methods depending on the criterion used for evaluation (Table 3).

**Table 2.** Results of the negative control experiments when the experimental and control arms are the same. MSE and MAE are respectively the mean squared error and the mean average error between the ATT estimation and the ground truth, which is null. Coverage is the percentage of confidence intervals that contain zero.

| | MSE($\times 1000$) | MAE($\times 100$) | C.I. width ($\times 1000$) | Coverage(%) |
| --- | --- | --- | --- | --- |
| Unadjusted | 8.73 | 7.62 | 24.9 | 78% (7/9) |
| PSM | 6.46 | 6.79 | 28.3 | 100% (9/9) |
| IPTW | 4.79 | 5.91 | 27.9 | 100% (9/9) |
| G-computation | **3.53** | **4.79** | 22.0 | 88% (8/9) |
| DDML | 4.72 | 5.70 | 28.9 | 100% (9/9) |

**Table 3.** Results of the RCT replication experiments. Pseudo MSE and MAE are respectively the pseudo mean squared error and the pseudo mean average error obtained by replacing the unknown ground truth with the RCT estimate. Estimate agreement is the percentage of RCT 95% confidence intervals that contain ATT estimation. Regulatory agreement is the percentage of time the cutoff *P* < 0.05 obtained from the non-randomized experiments agrees with the RCT result about *P* < 0.05.

| | Pseudo MSE($\times 1000$) | Pseudo MAE($\times 100$) | C.I. Width($\times 100$) | Estimate Ag. | Regulatory Ag. |
| --- | --- | --- | --- | --- | --- |
| Unadjusted | 7.94 | 7.30 | 25.1 | 84.2% (16/19) | 73.7% (14/19) |
| PSM | 4.51 | 6.15 | 29.0 | 89.5% (17/19) | 73.7% (14/19) |
| IPTW | 5.75 | 5.86 | 28.5 | 89.5% (17/19) | 78.9% (15/19) |
| G-computation | **3.26** | **4.68** | 25.9 | 100% (19/19) | 78.9% (15/19) |
| DDML | 4.70 | 5.60 | 31.3 | 100% (19/19) | 84.2% (16/19) |

**Figure 5.**
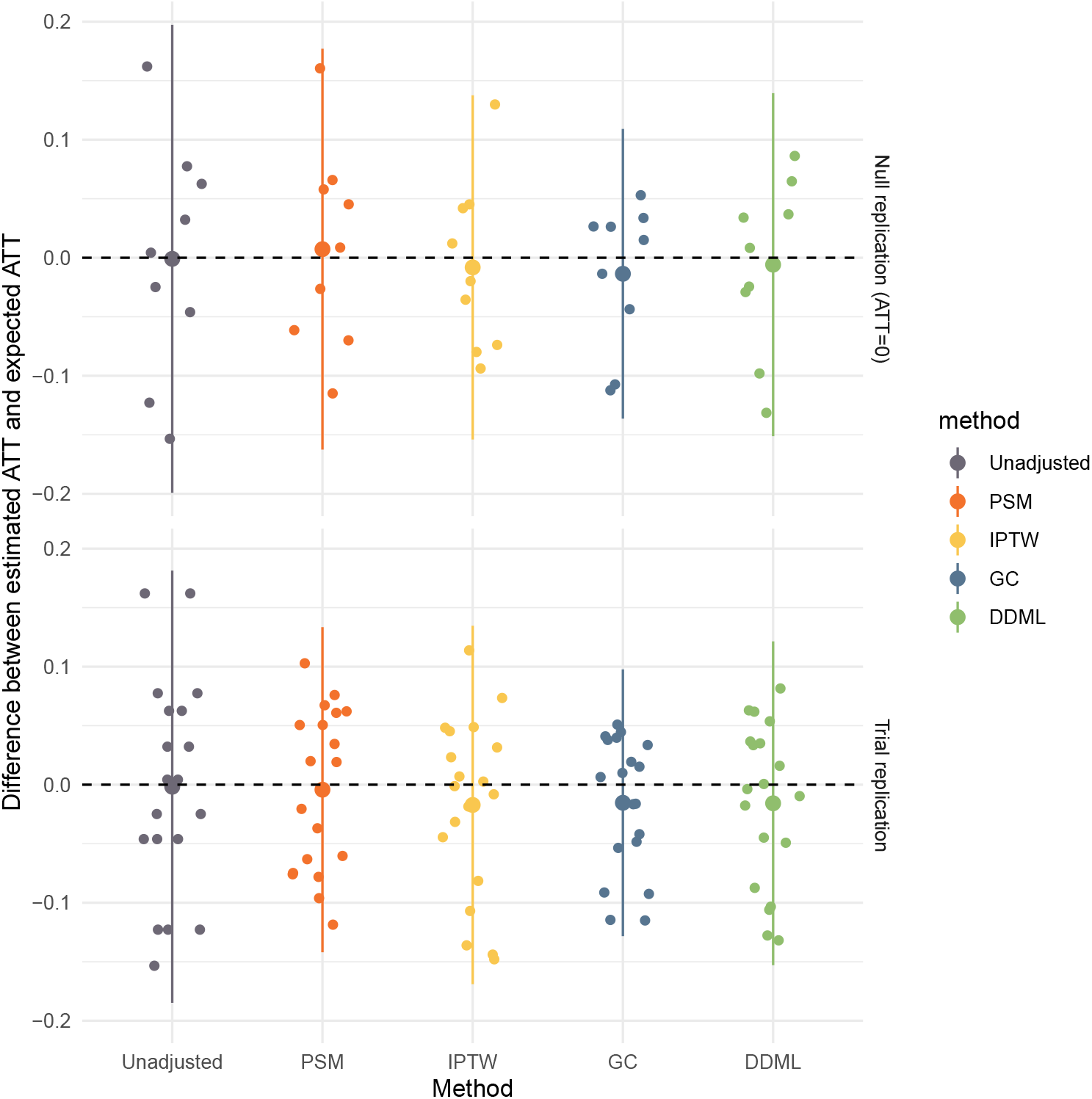
Results of the two replication experiments. Each point corresponds to an observational experiment. For the 9 null replication experiments, the expected target ATT is 0 and for the 19 RCT experiments, the expected target ATT is the RCT estimate. The larger point is the mean of the points and the bar extends to the mean plus or minus two times the standard deviation. For each method, the position on the x-axis does not matter and random perturbation on the x-axis is added to the points to allow optimal visualisation.

Width of confidence intervals also varies between methods (Tables 2 and 3). The G-computation method has the smallest width of C.I., the DDML methods has the largest width and the C.I. widths obtained with propensity-score methods are in between. The results mimic what is found at *n* = 250 for the synthetic simulations; the smallest width of C.I is found with G-computation and the largest one is obtained with DDML (Figure 4).

We also investigate coverage for the null replication (Table 2). The unadjusted method has the lowest coverage (7/9) whereas the propensity-score methods and DDML have complete coverage (9/9). The G-computation has intermediate coverage (8/9) reflecting its narrower confidence intervals.

In terms of estimate and regulatory agreement for the trial replication experiment, DDML has better agreement with trial results followed by G-computation (Table 3). However, differences between the two methods are small; there is regulatory agreement for 16 out of 19 trials with DDML whereas there is regulatory agreement for 15 out of 19 trials with the G-computation method. Agreement with trial results is inferior for propensity-score methods.

## Discussion

Based on both synthetic simulations and a replication study of completed randomized trials, we show that statistical methods based on outcome prediction models estimate treatment effect (ATT) more precisely than propensity-score methods, which confirms previous simulation results [19, 21]. Outcome prediction methods have correct type I errors while their power is generally greater than power of propensity score approaches. G-computation methods have increased power compared to propensity score approaches whatever the sample size. The results are more tempered for the DDML approach that explicitly accounts for the bias of machine learning models. For small sample sizes of *n* = 250 individuals, power of DDML can be reduced compared to propensity score methods whereas it is comparable to the power of G-computation for large sample size of *n* = 1000 patients.

There are marked differences between the results obtained with G-computation and DDML. As expected by construction of the DDML estimator, its bias is smaller than the bias of G-computation, which is in turn smaller than the bias of propensity score approaches. Another marked difference concerns the estimation of variance in order to compute credibility intervals. The sample splitting approach overestimates variance of DDML estimator. As a consequence, the widths of credibility intervals for DDML are increased that explains why type I errors are below the 5% threshold rate. DDML being conservative comes at a price of a 15 − 20% reduction of power compared to G-computation when the sample size is small (*n* = 250). By contrast G-computation is slightly too liberal; type I errors are between 5% and 10% and confidence intervals are the narrowest of all methods we considered. Propensity score methods have also increased type I error in some simulation setting which confirm previous simulation findings about excess of type I error of a propensity score matching method [46].

In practice, choosing between DDML and G-computation can be guided by at least two factors. The first factor is sample size as we found that DDML relative performance depends on sample size. In practice, sample sizes for external control arms can have different orders of magnitude ranging from dozens to thousands of patients [47]. In oncology, after application of inclusion and exclusion criteria, sample size can be smaller than *n* = 100 [48] where G-computation should be preferred, but can also exceed *n* = 500 where application of DDML can be preferred [49]. The second factor is the dimension of confounding covariates that is related to the risk of bias for machine learning estimator. When considering one or several dozens of confounding variable, risk of bias is small or moderate (for very small sample size) and G-computation can be considered. However, in future applications of external control arms, confounding variables can be high dimensional data such as as genomics, imaging, or electronic health record data. Observational experiments where high-dimensional confounders have been measured is emerging because of availability of electronic health record data and DDML can be relevant in this setting [50, 51]. When risk of bias exists because of overfitting, the DDML estimator should be preferred.

External control arm (ECA) analysis considerably reduces the risks of false positive errors of single arm-trial because it adjusts for the clinical profiles of patients [3]. However, ECA analyses, and more generally RWE analyses, do not fully reproduce results of randomized studies [49, 52, 53]. Therefore, it provides a valuable and less liberal estimation of efficacy than single arm studies [3] but it is not a substitute for large randomized studies. In this paper, we have shown that machine learning methods such as G-Computation and DDML, can improve external control arm analyses by increasing statistical power while preserving type I error.

## Data Availability

Data access should be requested to the Yale University Open Data Access (YODA) Project https://yoda.yale.edu/how-request-data

## Appendix

### Method to compute DDML estimator and its variance

To take into account the variability of the splitting procedure in the computation of the DDML estimator (equation (3)), the split is repeated *S* × *K* times by repeating *S K*-fold cross-validation procedures. For a cross-validation scheme, the aggregated estimator is the average of the estimators obtained using the *k*^*th*^-fold, *k* = 1, …, *K*, as the auxiliary dataset

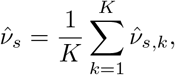

where *ν*_*s,k*_ is the ATT estimator 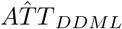 (Equation (3)) for the *s*^th^ cross-validation repetition, considering the *k*^th^ fold as the auxiliary dataset, and the remaining folds to train the two machine learning models. The overall estimate is obtained as

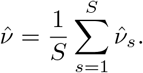

The variance of 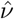 is estimated with

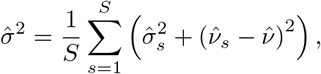

where

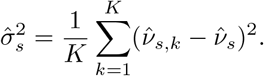

**Appendix Table 1.** Distribution of main variables used in the replication analysis

| Trials | NCT01137812 | NCT01106625 | NTC01106651 | NCT01106677 | NCT00968812 |
| --- | --- | --- | --- | --- | --- |
| Variables |  |  |  |  |  |
| Age | 56.86<br>(± 9.25) | 56.30<br>(± 9.33) | 63.55<br>(± 6.27) | 55.46<br>(± 9.33) | 55.95<br>(± 9.28) |
| Sex (Male) | 52.3% | 6.5% | 55% | 47% | 53% |
| HBA1C | 8.12<br>(± 0.92) | 8.12<br>(± 0.91) | 7.70<br>(± 0.78) | 7.94<br>(± 0.90) | 7.79<br>(± 0.78) |
| BMI | 33.0<br>(± 6.49) | 31.6<br>(± 6.76) | 31.6<br>(± 4.60) | 31.9<br>(± 6.10) | 31.1<br>(± 5.35) |
| LDL | 2.70<br>(± 6.49) | 2.54<br>(± 6.76) | 2.33<br>(± 4.60) | 2.78<br>(± 6.10) | 2.66<br>(± 5.35) |
| Blood Glucose Level | 12.62<br>(± 7.54) | 13.60<br>(± 6.97) | 10.60<br>(± 2.36) | 11.03<br>(± 13.29) | 8.83<br>(± 1.94) |

We always consider 3-fold cross validation. In the Monte Carlo simulations, we consider *S* = 20 repetitions, and in the internal replication, we consider *S* = 10 repetitions.

### List of the variable included in the propensity score/outcome model

Serum Albumin, Alkaline phosphatase, Alkaline transaminase, Aspartate transaminase, Basophils/Leukocytes, Biliburin, Blood Urea Nitrogen, Calcium, Cholesterol, Creatine Kinase, Chloride, Serum Creatinine, Eosinophils, Glomerular Filtration Rate Corrected, Gamma-Glutamyl Transferase, Blood sugar level, Plasma Glucose, Hemoglobin A1C, HDL Cholesterol, Hemoglobin, Potassium, LDL, Lymphocytes, Lymphocytes/Leukocytes, Magnesium, Neutrophil, Phosphate, Platelets, Protein, Sodium, Triglycerides, Diastolic Blood Pressure, Systolic Blood pressure, Pulse Rate, Weight, Age, Sex, Race Black or African American, Race other, Race white, Zone asia pacific, Zone central South America, Zone north America, Tabacco use, Concomitant medication diabetes, Previous concomitant medication anti-hyperglycemic, previous concomitant therapy

## Acknowledgements

This study, carried out under YODA Project #2019-4077, used data obtained from the Yale University Open Data Access Project, which has an agreement with JANSSEN RESEARCH & DEVELOPMENT, L.L.C.. The interpretation and reporting of research using this data are solely the responsibility of the authors and does not necessarily represent the official views of the Yale University Open Data Access Project or JANSSEN RESEARCH & DEVELOPMENT, L.L.C.

## Abbreviations

ATT: Average Treatment effect on the Treated
ATE: Average Treatment Effect
C.I.: Confidence Intervals
DDML: Doubly Debiased Machine Learning
ECA: External Control Arm Analysis
IPTW: Inverse Probability of Treatment Weighting
MAE: Mean Absolute Error
MSE: Mean Squared Error
PSM: Propensity Score Matching
RCT: Randomized Clinical Trial
YODA: Yale University Open Data Access

## Availability of data and materials

Data access should be requested to the Yale University Open Data Access (YODA) Project.

## Consent for publication

All contributing parties have consented for this work to be published.

## Authors’ contributions

G.W., M.B., N.L., and P.T. designed the study and experiments. M.H. and M.Z. gave feedback at various stages of the study. N.L. and P.T. performed experiments. M.B. wrote the manuscript that was revised by G.W., M.A., N.L. and P.T.

